# The construction and validation of sub-phenotype-specific genetic risk scores in systemic lupus erythematosus: a novel approach using large-scale biobank data

**DOI:** 10.1101/2025.03.11.25323758

**Authors:** Sarah Reid, Johanna K Sandling, Pascal Pucholt, Ahmed Sayadi, Martina Frodlund, Karoline Lerang, Iva Gunnarsson, Andreas Jönsen, Ann-Christine Syvänen, Øyvind Molberg, Solbritt Rantapää-Dahlqvist, Anna Rudin, Christopher Sjöwall, Elisabet Svenungsson, Anders A Bengtsson, Lars Rönnblom, Dag Leonard

## Abstract

Systemic lupus erythematosus (SLE) is an autoimmune disease with a heterogenous clinical picture. This study aimed to link genetic SLE predisposition with relevant clinical manifestations using a two-step approach. First, we identified datasets best corresponding to the 11 American College of Rheumatology 1982 (ACR-82) classification criteria for SLE using an ICD-10 code-based search in a large, public database (FinnGen consortium). Mendelian Randomization analysis of these datasets linked genetic SLE predisposition to several SLE-like manifestations: rosacea, OR 1.09(1.03–1.16), polyarthropathies, OR 1.10(1.06–1.14), pleural effusions, OR 1.09(1.04–1.14), and hemolytic anemia, OR 1.32(1.10–1.58). Second, validation was conducted in a clinical SLE cohort comprising 1,487 genotyped Scandinavian patients with detailed medical records. Based on the public datasets, genetic risk scores (GRS) for each relevant manifestation were constructed for each patient. Associations between each GRS and the corresponding ACR-82 criterion were evaluated using sex- and disease duration-adjusted logistic regression. Five of the 11 ACR-82 criteria were associated with their corresponding GRS: arthritis, OR 1.15(1.02–1.31), nephritis, OR 1.15(1.04–1.29), neurology, OR 1.24(1.04–1.47), hematology, OR 1.12(1.00–1.24), and immunology, OR 1.37(1.22–1.56), indicating that our method of using publicly available datasets to construct manifestation-specific GRSs may be useful in predicting SLE outcomes.

## Introduction

Systemic lupus erythematosus (SLE) is a chronic autoimmune disease with a mortality rate exceeding that of the normal population.[1, 2] Its highly heterogeneous clinical presentation and unpredictable disease course often make early distinction between mild and severe cases challenging, with current clinical methods having a very limited ability to predict future manifestations or complications.[3–5] To optimize treatment strategies for the individual patient, new tools for risk stratification are essential. Genetic risk scores (GRSs), calculated by summarizing the effect of several genetic variants, can provide insights into the cumulative genetic predisposition to SLE. In recent studies, strong associations have been demonstrated between a high GRS for the disease and the development of cardiovascular and renal manifestations, including a doubled risk of nephritis and more-than five-fold risk of end-stage renal disease in patients with a high GRS compared to those in the low quartile.[6–8] These findings suggest that within the genetic risk of SLE, there are sub-groups of genetic loci associated with individual disease manifestations. Identifying these loci could potentially increase the predictive ability of GRSs further, offering a method of discriminating between mild and severe SLE early in the disease course. However, due to the small effect sizes of each individual locus and small cohort sizes due to the low disease prevalence, significant progress in identifying individual SNP-sub-phenotype associations and constructing phenotype-specific GRSs has so far been limited.[8]

The aim of this study was to identify groups of established SLE risk SNPs that are associated with typical sub-phenotypes of SLE, such as arthritis, serositis, nephritis, and hematologic and immunologic disturbances.[9] First, Mendelian Randomization (MR) analysis was employed to explore the impact of a high genetic SLE predisposition on a number of SLE-like manifestations in a large population, predominantly consisting of individuals without SLE. Second, a clinical cohort of Scandinavian patients with SLE was investigated to determine the relevance of the associations found in the non-SLE population for predicting the occurrence of each ACR-82 classification criteria in SLE.

## Methods

### Study design

This study includes analyses performed both on summary level data from the FinnGen Consortium, and on individual patients with SLE (Figure 1). The FinnGen research project is based on samples from Finnish biobanks and data from national health registers, and data access is available through the FinnGen consortium.[10] The University of Helsinki is responsible for the FinnGen research project and is the official data controller of the study, and the Coordinating Ethics Committee of the Helsinki and Uusimaa Hospital District have provided ethical approval of the project.[10]

**Figure 1.**
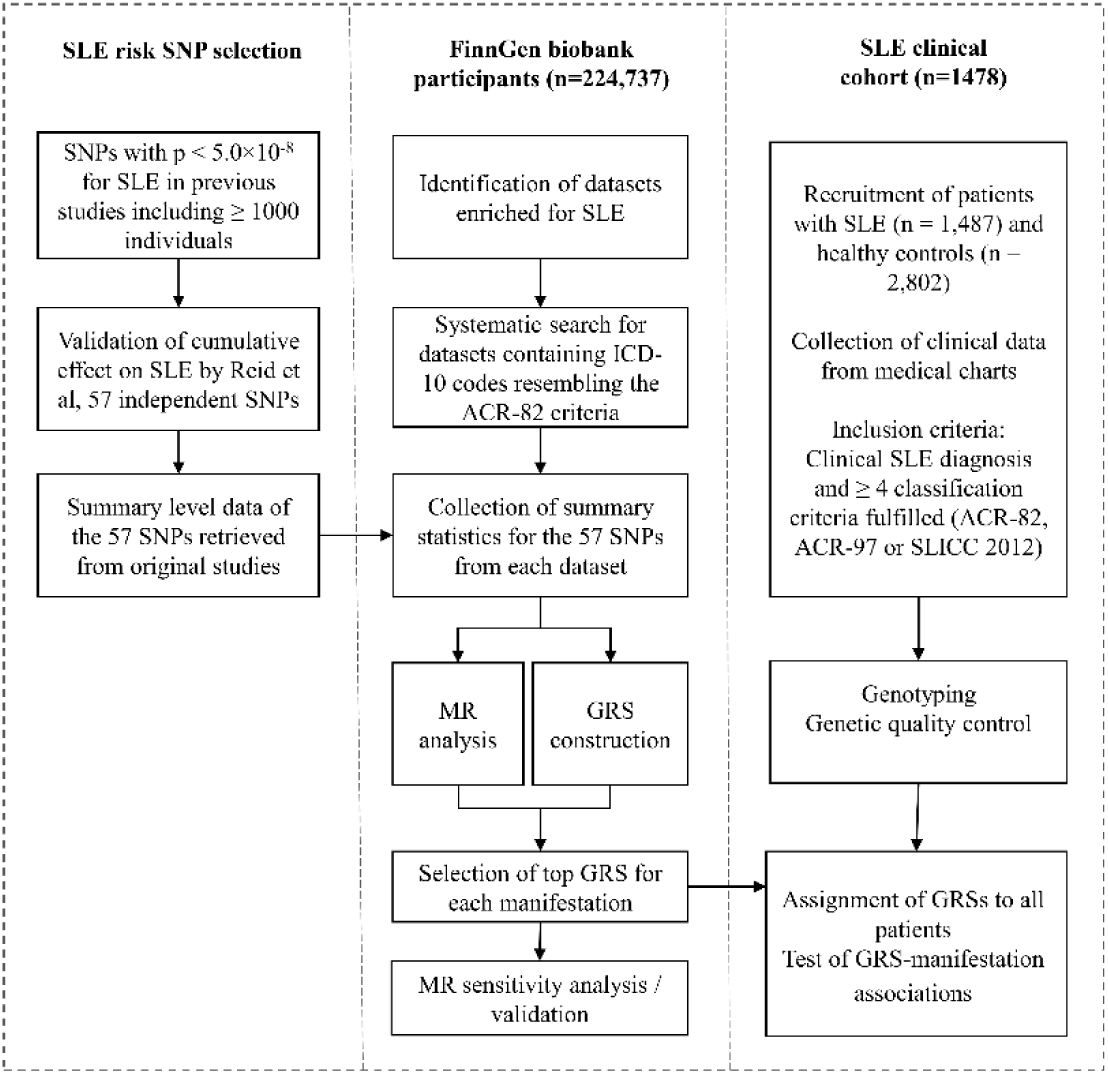
Method overview. ICD-10: International Classification of Diseases-10.[11] ACR: American College of Rheumatology.[9, 12] MR: Mendelian randomization. SLICC: Systemic Lupus Collaborating Clinics.[13] GRS: genetic risk score.

### Mendelian Randomization analysis and data sources

MR is a genetics-based instrumental variable (IV) approach, relying on the random assignment of genetic variants at conception, to investigate the causal effect of an exposure on an outcome.[14, 15] In the present study, genes associated with SLE development in the European population were employed as IVs, and their effect on a number of SLE-like manifestations was evaluated. We have previously constructed a GRS for SLE which, when validated in a large cohort of over 5,000 European patients with SLE and 9,000 healthy controls, displayed one of the highest predictive accuracies for SLE development so far demonstrated (area under the receiver operating characteristics curve 0.71 in the discovery cohort).[7] The GRS consisted of 57 independent (r^2^<0.2) SLE risk SNPs which in turn have displayed individual association (p<5×10^-8^) with SLE in previous studies performed on the European population, including at least 1,000 individuals.[16–31] The individual and cumulative effects of these SNPs on SLE are thus well-explored and meet the MR relevance assumption, and the current study therefore employs these 57 SNPs as IVs.[14, 15] For each of these SNPs, the original genome-wide association study (GWAS) was used to retrieve summary level statistics, including information on the risk/protective allele, effect sizes, standard errors or confidence intervals (CIs), and p-values (Table 1)

**Table 1.**
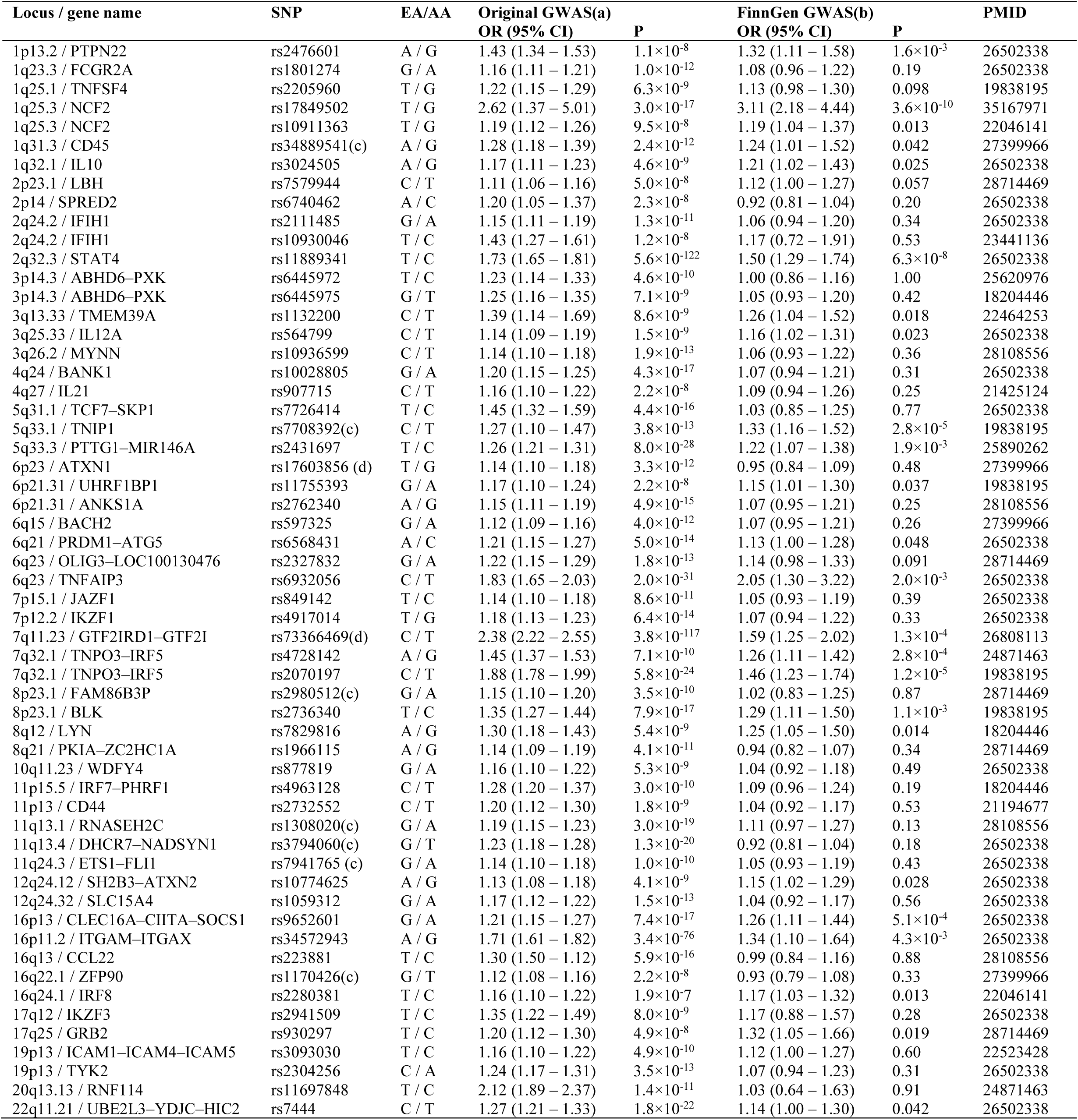
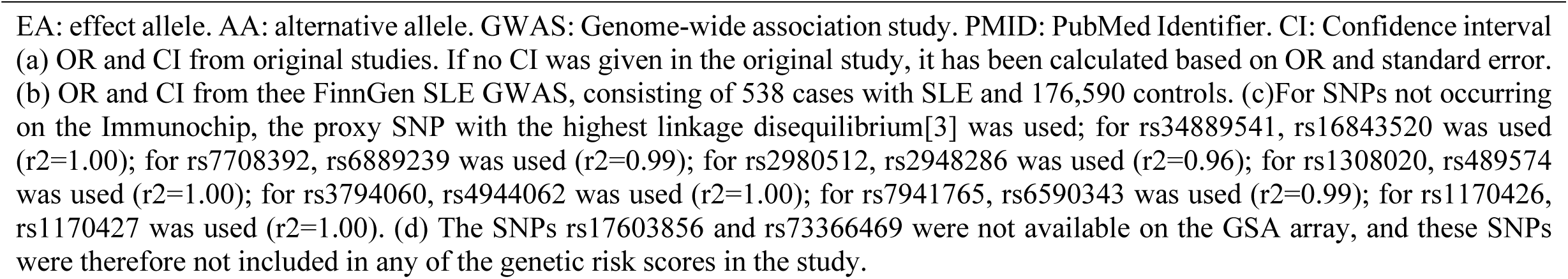
The 57 SNPs with previously established association with SLE development.

The outcomes of interest in this study were the manifestations in the FinnGen database best corresponding to each of the ACR-82 classification criteria.[9] Data from the FinnGen consortium (Data Freeze 5, 2021) was used to identify appropriate outcome datasets.[10] Information about available datasets was obtained from the Risteys portal (2021).[32] The search was limited to datasets including International Classification of Diseases (ICD)-10 codes corresponding to *diseases of the skin and subcutaneous tissue* (L00–L99), *diseases of oral cavity, salivary glands and jaws* (K00–K14), *diseases of the musculoskeletal system and connective tissue* (M00–M99), *other forms of heart disease* (I30–I52), *other diseases of pleura* (J90–J94), *glomerular diseases* (N00–N08), *diseases of the nervous system* (G00–G99), *diseases of the blood and blood–forming organs and certain disorders involving the immune mechanism* (D50–D89), and *abnormal findings on examination of blood, without diagnosis* (R70–R79).[11] To ensure relevance for SLE of selected datasets, only datasets including a more-than-two-fold higher prevalence of SLE compared with the entire FinnGen database were selected. When two datasets contained largely overlapping ICD-10 codes, the largest dataset was selected. In total, 30 datasets were identified (Figure 2). In addition, data from a FinnGen SLE GWAS (ICD-10 code M32) performed on 538 patients and 213,145 controls was retrieved, Figure 2.[32]

**Figure 2.**
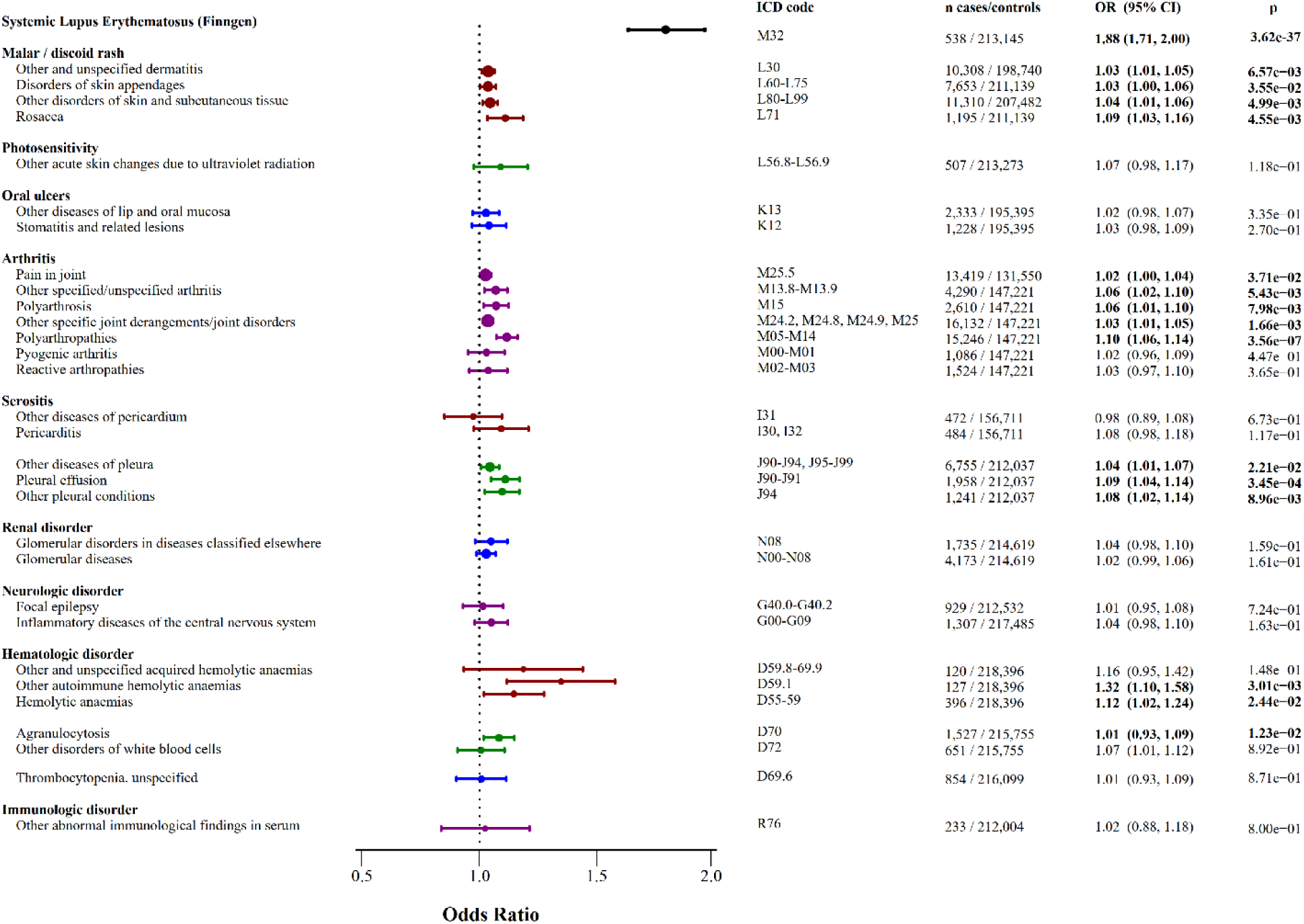
Results from the Mendelian Randomization (MR) analyses. The SLE dataset and 30 manifestations best corresponding to each of the ACR-82 classification criteria,[9] were selected from the FinnGen database[32] and analyzed through MR using the inverse variance weighted (IVW) method.

Summary statistics for each of the 57 SLE risk SNPs were retrieved for each dataset using the TwoSampleMR R package. The exposure dataset was subsequently harmonized with each of the outcome datasets to ensure that the measured effect of a particular SNP on both the exposure and the outcome corresponded to the same allele.

### SLE clinical cohort and healthy controls

The clinical cohort included 1,487 patients with SLE from Sweden and Norway. All subjects fulfilled ≥ 4 ACR-82, ACR-97 or the SLICC-2012 classification criteria for SLE and were of European descent. Clinical data, including the ACR-82 classification criteria, disease duration, sex and age at follow-up, was collected from medical records. Clinical characteristics of the cohort are described in Table 2. Control individuals were healthy blood donors from Uppsala (Uppsala Bioresource) and Lund or population-based controls from Stockholm and the medical biobank in Umeå, which contains population-based samples from the Västerbotten country, Sweden. All patients in the clinical SLE cohort provided their informed consent, and the study protocol was approved by the Regional Ethical Review Board, Uppsala (DNR 2009/013 and 2020– 05065) and the local ethics committees.

**Table 2.**
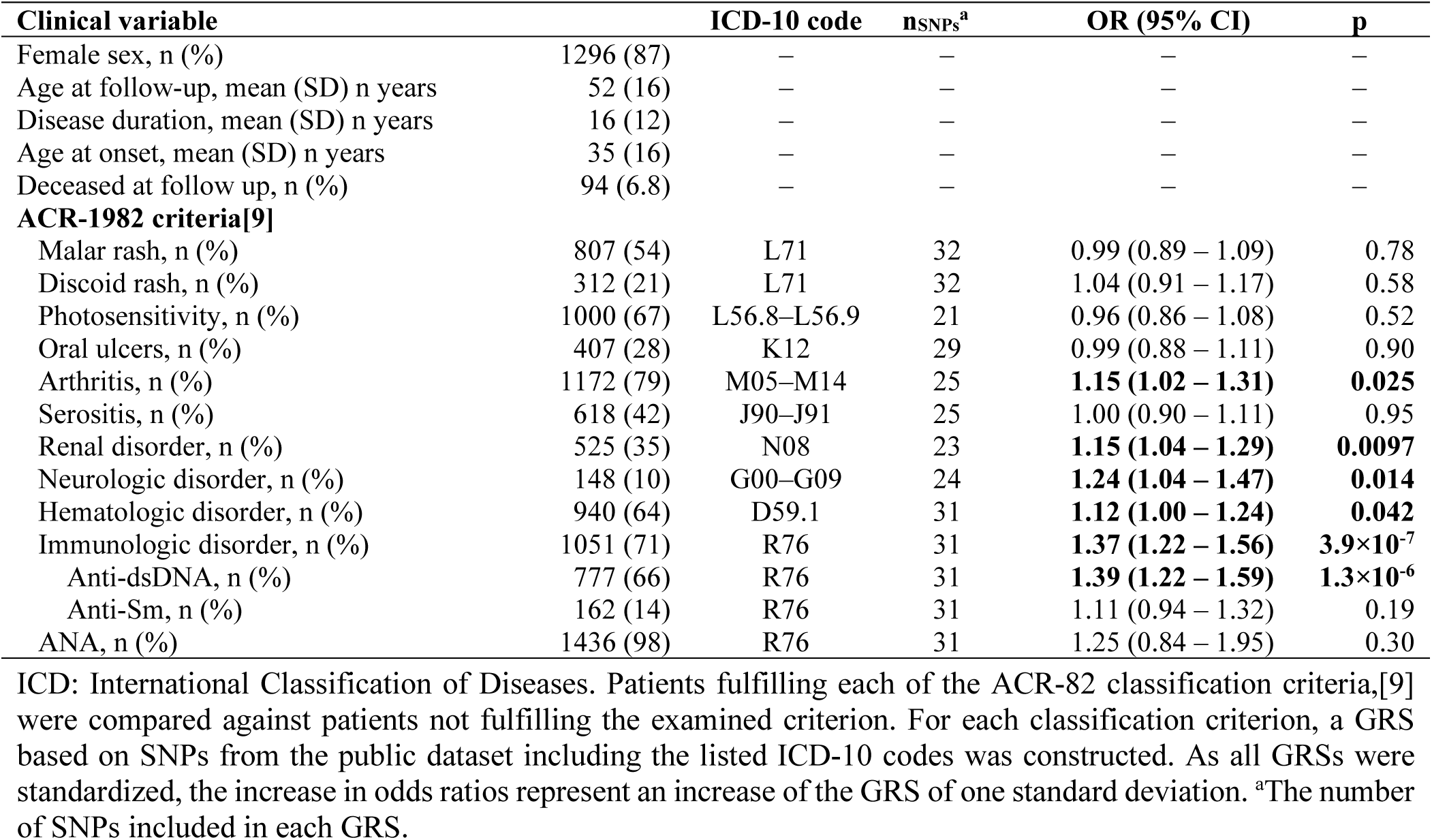
Validation of the genetic risk scores (GRSs) for disease sub-phenotypes in patients with SLE (n=1487).

| Clinical variable |  | ICD-10 code | nSNPs<sup>a</sup> | OR (95% CI) | p |
| --- | --- | --- | --- | --- | --- |
| Female sex, n (%) | 1296 (87) | — | — | — | — |
| Age at follow-up, mean (SD) n years | 52 (16) | — | — | — | — |
| Disease duration, mean (SD) n years | 16 (12) | — | — | — | — |
| Age at onset, mean (SD) n years | 35 (16) | — | — | — | — |
| Deceased at follow up, n (%) | 94 (6.8) | — | — | — | — |
| **ACR-1982 criteria[9]** |  |  |  |  |  |
| Malar rash, n (%) | 807 (54) | L71 | 32 | 0.99 (0.89 – 1.09) | 0.78 |
| Discoid rash, n (%) | 312 (21) | L71 | 32 | 1.04 (0.91 – 1.17) | 0.58 |
| Photosensitivity, n (%) | 1000 (67) | L56.8–L56.9 | 21 | 0.96 (0.86 – 1.08) | 0.52 |
| Oral ulcers, n (%) | 407 (28) | K12 | 29 | 0.99 (0.88 – 1.11) | 0.90 |
| Arthritis, n (%) | 1172 (79) | M05–M14 | 25 | **1.15 (1.02 – 1.31)** | **0.025** |
| Serositis, n (%) | 618 (42) | J90–J91 | 25 | 1.00 (0.90 – 1.11) | 0.95 |
| Renal disorder, n (%) | 525 (35) | N08 | 23 | **1.15 (1.04 – 1.29)** | **0.0097** |
| Neurologic disorder, n (%) | 148 (10) | G00–G09 | 24 | **1.24 (1.04 – 1.47)** | **0.014** |
| Hematologic disorder, n (%) | 940 (64) | D59.1 | 31 | **1.12 (1.00 – 1.24)** | **0.042** |
| Immunologic disorder, n (%) | 1051 (71) | R76 | 31 | **1.37 (1.22 – 1.56)** | **3.9×10<sup>-7</sup>** |
| Anti-dsDNA, n (%) | 777 (66) | R76 | 31 | **1.39 (1.22 – 1.59)** | **1.3×10<sup>-6</sup>** |
| Anti-Sm, n (%) | 162 (14) | R76 | 31 | 1.11 (0.94 – 1.32) | 0.19 |
| ANA, n (%) | 1436 (98) | R76 | 31 | 1.25 (0.84 – 1.95) | 0.30 |

ICD: International Classification of Diseases. Patients fulfilling each of the ACR-82 classification criteria,[9] were compared against patients not fulfilling the examined criterion. For each classification criterion, a GRS based on SNPs from the public dataset including the listed ICD-10 codes was constructed. As all GRSs were standardized, the increase in odds ratios represent an increase of the GRS of one standard deviation. <sup>a</sup>The number of SNPs included in each GRS.

### Genotyping and construction of genetic risk scores

1001 of the SLE patients and the 2802 controls had previously been genotyped using the Illumina 200K Immunochip SNP array by the SNP&SEQ Technology platform at Science for Life Laboratory in Uppsala, Sweden. Quality control procedures have previously been described.[7] An additional 486 unique SLE patients genotyped using the Global Screening array by the SNP&SEQ Technology platform at Science for Life Laboratory in Uppsala, Sweden, were included in the study. For quality control (QC) procedures and selection of patients, see supplementary methods. Of the 57 SLE risk SNPs, 55 were available on the GSA array (Table 1). Data for each of the 55 SNPs was available for all included individuals.

For each of the nine manifestations investigated in the MR analysis, the most significant dataset was selected for GRS construction. For each of the 55 SNPs where the direction of the effect size for the examined outcome matched that of SLE, the natural logarithm of the OR for the outcome was multiplied by the number of risk alleles in each individual. The sum of all products for each patient was defined as the GRS. To facilitate interpretation and comparisons of effect sizes, all GRSs were standardized by subtraction of the mean value and division by one standard deviation.

### Statistical analysis

Using summary statistics for the 57 SLE SNPs as described above, MR was conducted for each of the 30 outcome datasets using the TwoSampleMR package in R software 4.0.2 (R Foundation for Statistical Computing, Vienna, Austria). The inverse variance weighted (IVW) method was employed as the main method to test for a significant association between the genetic risk of SLE and the outcome variables. One dataset per ACR-82 classification criteria was subsequently selected for further analyses and construction of GRSs. Four additional MR methods, including the MR Egger method, simple and weighted median methods and a sign concordance test, were used for additional sensitivity analyses on these datasets, and pleiotropic effects were assessed by evaluating the extent to which the MR-Egger intercept was non-zero and by assessing the Q statistics for heterogeneity with respect to IVW. To validate each of the nine constructed GRSs in the clinical cohort of SLE patients, logistic regression models were used. All analyses were adjusted for sex, and all case-case analyses were adjusted also for disease duration. In both the MR analyses and the analysis of the clinical cohort, p-values<0.05 were considered statistically significant.

## Results

### The SLE genetic risk and occurrence of SLE in the FinnGen biobank

In the FinnGen SLE dataset, 538 individuals (0.25%) had been diagnosed with SLE (ICD-10 code M32), with a mean age at onset of 46 years, a female:male ratio of 5:1, and a case fatality rate of 3.1%.[32] As the original GWASs leading to the identification of the 57 SLE risk SNPs were based on non-Finnish Europeans, which genetically display slight differences compared with the Finnish population,[33] an initial validation of these SNPs for SLE was performed (Figure 2). Comparing the prevalence of each SNP between the 538 cases and 213,145 controls, 24 of the 57 SNPs were significantly associated with SLE, with several well-known SNPs showing strong association in the cohort (Figure 3A) and one SNP displaying association below the GWAS significance threshold (rs17849502 (*NCF2*), p=3.6×10^-10^, Table 1). In the MR analysis, the combined effect of the 57 SNPs was a strong predictor of SLE development (OR 1.88 (1.71–2.00), p=3.6×10^-37^, Figure 2). The risk allele frequencies of each SNP in the healthy controls of the SLE FinnGen GWAS corresponded well to those of the Swedish controls (r=0.99, p<2.2×10^-16^), with no identified outliers (Figure 3B).

**Figure 3.**
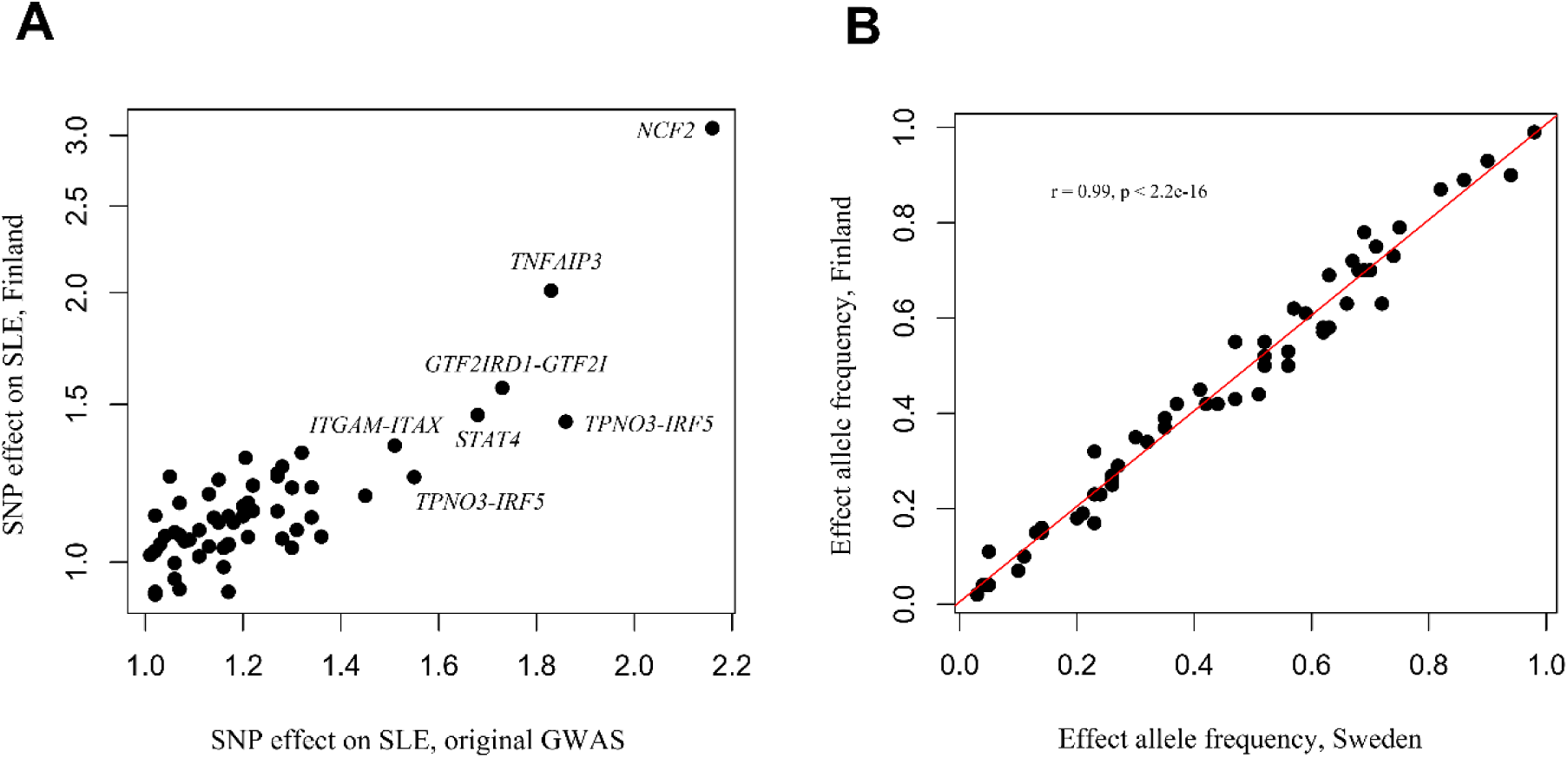
The effect of the 57 SLE risk SNPs in the FinnGen biobank. A) For each of the 57 SNPs included in this study, the odds ratios (ORs) for SLE reported in the original genome-wide association study (GWAS) leading to identification of the SNP, was plotted against the OR for SLE in the FinnGen biobank, based on a GWAS dataset including 538 SLE cases and 213,145 controls. B) The effect allele frequency of each of the 57 SNPs in the 2 802 Swedish healthy controls included in this study, was compared with the 213,145 controls included the FinnGen SLE dataset.

### The SLE genetic risk and occurrence of SLE-like manifestations in the FinnGen biobank

In the MR analysis of the public datasets, individuals with a stronger genetic predisposition to SLE were more likely to be diagnosed with several of the examined SLE-like manifestations (Figure 2). For example, all four skin manifestation datasets exhibited a significant increase in the likelihood of diagnosis with an increasing predisposition to SLE, with the strongest skin-related association found for rosacea (OR 1.09 (1.03–1.16), p=0.0046, Figure 2). Unlike the skin datasets, neither of the datasets for photosensitivity or oral ulcers displayed an association with the SLE genetic risk (Figure 2).

Seven datasets related to arthritis and arthropathies were examined, of which five exhibited a significant association with a high genetic SLE predisposition (Figure 2). The strongest association was found for polyarthropathies (OR 1.10 (1.06–1.14), p=3.6×10^-7^), whereas the two non-significant findings were for reactive arthropathies (p=0.37) and pyogenic arthritis (p=0.45), suggesting the genetic background differs for these manifestations.

Regarding risk for serositis, two datasets for pericarditis and three for pleural conditions were analyzed. A higher genetic predisposition to SLE was found to increase the risk of diagnosis in all three pleural manifestation datasets, with the strongest association identified for pleural effusion (OR 1.09 (1.04–1.14), p=0.00035, Figure 2). In contrast, the SLE genetic risk did not significantly increase the risk of pericarditis (Figure 2).

In terms of hematological manifestations, the SLE genetic risk was found to increase the risk of hemolytic anemia in two of the three examined datasets. The evidence of an association with leukopenia was weaker, with only one of the two datasets exhibiting a significant association, while no association was found in the thrombocytopenia dataset (Figure 2). For immunology, one dataset was examined, which included cases with raised antibody titers, a false-positive test for syphilis or other specified or unspecified immunological findings in serum. No statistically significant association with the SLE genetic risk was found for this dataset (Figure 2).

To test the robustness of the MR analyses, sensitivity analyses were performed using four additional MR methods (Supplementary Table 1). Overall, these analyses generated results consistent with the initial IVW method, with the four previously significant manifestations displaying similar effect sizes and most results remaining significant or close to significant, Supplementary Table 3. Employing the MR Egger test of intercept, no evidence of pleiotropy was identified in any of the datasets; however, the arthritis dataset displayed a clear sign of heterogeneity (p=8.0×10^-20^), indicating pleiotropic effects of individual SNPs (Supplementary Table 3). To ensure that the selection of datasets including an overlap with the FinnGen SLE dataset did not result in bias in the MR analyses, the correlation between the IVW results and the case overlap of all 30 datasets was assessed. On average, the overlap of the FinnGen SLE dataset with the 30 outcome datasets was 1.1% (range: 0.52–2.77%), with no evidence of a correlation between the case overlap and the p-values (r=-0.02, p=0.91) or ORs (r=0.0043, p=0.85) from the analyses.

For further analysis, the most strongly associated dataset for each of the ACR-82 criteria was selected. In these datasets, two SNPs displayed individual GWAS significance for any manifestation (rs2476601 (*PTPN22*) and rs10774625 (*SH2B3–ATXN2*), both for arthritis, Supplementary Table 2).

### Clinical SLE cohort validation

In the clinical cohort, the mean age at follow-up was 52 years, the mean disease duration was 16 years and the mean age at disease onset was 35 years (Table 2). Initially, each of the nine constructed manifestation GRSs was compared for patients who met each of the ACR-82 criteria against healthy controls, and all GRSs were found to be highly associated with their respective outcomes (all p<10×10^-16^, Supplementary Table 3). However, as these results may reflect the genetic risk of SLE as a whole rather than each individual manifestation, a comparison of patients with and without each manifestation was subsequently carried out. In these case-case analyses, the GRSs based on polyarthropathies, glomerulonephritis, neuroinflammation, hemolytic anemia and immunologic disturbances were found to be associated with the ACR-82 manifestations arthritis (OR 1.15 (1.02–1.31), p=0.025), renal disorder (OR 1.15 (1.04–1.29), p=0.0097), neurologic disorder (OR 1.24 (1.04–1.47), p=0.014), hematologic disorder (OR 1.12 (1.00–1.24), p=0.042) and immunologic disorder (OR 1.37 (1.22–1.56), p=3.9×10^-7^), respectively, Table 2. In contrast, there was no evidence of an association between the GRSs based on the datasets for rosacea, photosensitivity, stomatitis or pleural effusions and their corresponding ACR-82 criteria, Table 2. Analysis of SNP overlap between the different GRSs revealed moderate correlations between the scores (mean r=0.47, Supplementary Figure 1), with several SNPs contributing to multiple GRSs (Supplementary Table 2). The overlap was particularly notable for arthritis and hematologic disorder (r=0.69) and photosensitivity and neurologic disorder (r=0.67), respectively (Supplementary Figure 1), indicating shared genetic risk factors and potentially common etiological pathways between these manifestations.

## Discussion

This study aimed to explore the association between a high genetic risk of SLE and typical manifestations of the disease. We employed MR analysis to investigate the effect of 57 well-established SLE risk SNPs on a number of SLE-like manifestations in the FinnGen biobank, and then constructed GRSs which were validated in a large clinical cohort of patients with SLE.

An important finding of this study is that in the clinical cohort, we succeeded in constructing a GRS related to the manifestation for five of the nine selected FinnGen datasets. While the cumulative genetic risk of SLE is previously known to be associated with a smaller number of SLE sub-phenotypes, specifically nephritis and anti-dsDNA antibodies, we demonstrate a new approach to detect the presence of sub-groups of SNPs within the known cumulative genetic SLE risk.[6–8] These sub-groups were associated with several additional ACR-82 criteria, specifically arthritis, neurologic disorder, and hematologic disorder.

Prior studies have faced challenges in developing GRSs based on SNPs associated with SLE sub-phenotypes, with one of the most comprehensive studies, including over 3,000 patients with SLE in the discovery cohort, found to be underpowered to detect lupus nephritis SNPs at GWAS significance or to construct a GRS for the manifestation.[6–8] With the increasing number of individuals being included in large biobanks, our approach of not limiting the discovery analysis to patients with SLE, may overcome the challenges brought on by the rarity of the disease and small effect sizes of each genetic locus.

The MR analysis revealed that a high genetic risk of SLE increased the risk of skin manifestations, polyarthropathies, pleural effusion and hemolytic anemia in the FinnGen cohort. Some of these associations may reflect a shared genetic background and shared biological mechanisms with SLE. It is known that in certain individuals, years can pass between the onset of the first SLE manifestation and the time of clinical diagnosis and, although the prevalence of patients diagnosed with SLE was low (<3%) in all examined datasets, we speculate that the prevalence of cases with early or “incomplete” SLE may be higher.[34]

For other associations revealed by the MR analysis, the association may not be directly linked to SLE but could be due to pleiotropic effects of the included IVs. While our analyses did not find evidence of pleiotropy for any of the examined manifestations, we did find evidence of strong heterogeneity among individual SNPs in the polyarthropathies dataset, likely reflecting the shared genetic background between SLE and other rheumatic diseases associated with arthritis.[35] This observation may be important for the development of refined predictive models for clinical outcomes in SLE, where the inclusion of variants associated with genetically similar diseases may lead to improved accuracy.

In addition to a shared etiological mechanism, a diagnostic overlap between SLE and other diseases may have influenced some of our findings. For example, the HLA DRB1*03:01 allele is amongst the strongest genetic associations identified both for rosacea and SLE, indicating a possible shared genetic background for the diseases.[23, 36] The IVs included in the present study were all located outside the HLA region, and the association found between SLE and rosacea may thus be an additional indication of a connection in the etiology. However, as the appearance of an SLE malar rash and rosacea can be very similar, this result could also be due to the rosacea dataset containing a number of misdiagnosed patients with early or incomplete SLE.

While we found significant associations in several of the examined datasets, other manifestations displayed no significant association with the genetic risk of SLE. A possible reason for this is the discrepancy between the examined ICD-10 codes and the corresponding ACR-82 classification criteria. For example, the dataset selected to represent the ACR-82 nephritis criterion included glomerular disorders secondary to diseases like amyloidosis, and multiple myeloma, which likely have a different etiology, highlighting the necessity for greater granularity in diagnostic codes.[11] Moreover, some manifestations, such as skin changes due to UV radiation, may have mainly non-genetic causes, as supported by findings of a negative association between genetic risk of SLE and photosensitivity in a previous study.[7] In addition, the statistical power was limited by the small number of cases in the datasets for pericarditis (<500 cases) and immunological disturbances in serum (233 cases).

In addition to evaluating the SLE genetic risk and the risk of clinical manifestations, this is the first study to test the cumulative effect of a number of well-established SLE risk SNPs on the risk of SLE in the Finnish population. While the original FinnGen GWAS only identified three SNPs associated with SLE at GWAS significance, our results indicate a strong correlation between the effect sizes reported in previous studies on non-Finnish Europeans, and effect sizes in the Finnish population, with almost half of the individual SNPs reaching statistical significance in the present study.[10, 32] Our results thus indicate a large overlap in the genetic etiology of SLE between non-Finnish Europeans and the Finnish population. The reason for the low number of SNPs associated at GWAS significance in the dataset may consequently be due to low statistical power, or to identification of patients through ICD-10 codes rather than through assessment by a rheumatologist.

This study has several strengths, including the use of a large number of well-established SLE risk SNPs and a well-characterized clinical cohort of nearly 1,500 individuals with SLE. However, there are also some limitations to consider. Firstly, while MR is a useful tool for causal inference, all assumptions of the model cannot be directly tested for, and we therefore suggest that our results from the MR analyses should be interpreted as associations rather than causal relationships. Secondly, whilst the majority of the 57 SNPs used as IVs in this study were originally discovered in independent GWASs, some previous GWASs on SLE have included Swedish patients with SLE, which may lead to overestimation of the effect of these SNPs on SLE in the clinical SLE cohort. Lastly, the small influence of most of the GRSs on the ACR-82 criteria may suggest that only part of the genetic risk of each SLE manifestation can be captured using associations based on a non-SLE population. Moreover, the fact that each patient with SLE displays multiple manifestations of the disease adds additional complexity to our analyses and requires consideration of overlapping genetic factors involved in the manifestations. Due to the heterogeneity of SLE, the overlap between disease sub-phenotypes and the likely influence of both SLE-related and general genetic risk factors on disease severity, future studies should focus on developing more refined models that incorporate these overlapping genetic risk factors, as well as both SLE-specific and SLE-nonspecific genetic markers, to enhance the predictive power and clinical utility of GRSs.

In conclusion, this study demonstrates an association between a high genetic predisposition to SLE and several SLE-like manifestations. We have further shown that the approach of combining large biobank datasets with analysis of a well-characterized clinical SLE cohort may be useful in identifying part of the genetic etiology of specific SLE manifestations.

## Acknowledgements

We want to acknowledge the participants and investigators of the FinnGen study, the patients with SLE included in our clinical cohort, and the individuals who have participated as healthy controls in the study.

## Contributors

SR and DL designed the study. SR, MF, KL, IG, AJ, ØM, SRD, AR, CS, ES, AAB, LR and DL were involved in the collection of clinical data. JKS, PP, AS and ACS were involved in the genotyping and/or genetic quality control procedures. SR performed the review of public dataset and all statistical analyses. SR and DL analyzed the data. All authors revised the manuscript critically for important intellectual content and approved the final version of the manuscript

## Ethics approvals

The protocol for this study was approved by the Regional Ethical Review Board, Uppsala (DNR 2009/013 and 2020–05065) and the local ethics committees. Coordinating Ethics Committee of the Helsinki and Uusimaa Hospital District have provided ethical approval for the FinnGen project.

## Additional information

CS reports the following competing interests: Bristol-Myers Squibb(BMS) (employee). LR reports the following competing interests: Ampel Biosolutions (Advisor or Review Panel Member); AstraZeneca (Advisor or Review Panel Member, Speaker/Honoraria); Bayer (Consultant); BMS (Advisor or Review Panel Member); UCB (Advisor or Review Panel Member). DL reports the following competing interests: AstraZeneca (Advisor or Review Panel Member).

## Data availability

All public datasets used in this study are available for download from the FinnGen consortium (https://r5.finngen.fi/). As the data from the clinical cohort includes detailed genetic and clinical information, it cannot be openly shared. However, summary-level data and certain aggregated results may be made available upon reasonable request to qualified researchers, provided that the request complies with the ethical approvals and data sharing policies of the relevant institutions. Please contact the corresponding author for further information.

