## Supplementary file for "The construction and validation of sub-phenotype-specific genetic risk scores in systemic lupus erythematosus: a novel approach using large-scale biobank data"

**Supplementary methods**

**Genetic quality control procedures**

Of the 1 487 patients and the 2 802 control individuals included in this study, 1 001 patients had previously been genotyped using the Illumina 200K Immunochip SNP array by the SNP&SEQ Technology platform at Science for Life Laboratory in Uppsala, Sweden. Quality control procedures, as described previously, excluded individuals and SNPs with low call rates, those with abnormal heterozygosity, incorrect gender labels, and close relatives.[1] Principal components analysis (PCA) was utilized alongside the 1000 Genomes Project to remove outliers, and SNPs with minor allele frequency (MAF) <1% or significant deviation from Hardy-Weinberg equilibrium (HWE) in controls were also excluded.[1]

In addition, 1760 patients with SLE were genotyped using the Global Screening array (GSA) by the SNP&SEQ Technology platform at Science for Life Laboratory in Uppsala, Sweden. Samples with call rate below 100% and SNPs with call rate below 80% were excluded.[2] The HapMap project data was included in a principal component analysis (PCA) and utilized to remove samples more than 5 standard deviations (SD) from the European populations. To identify genetic relatedness between samples, plink was used to estimate the pairwise identity by descent (IBD). Samples with an IBD higher than 0.1875 were removed. Samples with heterozygosity rates more than 5 SD from the mean were excluded. A check for mislabeled gender was performed using Wright's inbreeding coefficient F, calculated from X chromosome data. Annotated females with F higher than 0.2 or annotated males with F lower than 0.8 were excluded. SNPs with a minor allele frequency (MAF) below 0.01 or which failed the Hardy–Weinberg equilibrium test (P–value $<1\times10^{-4}$), were excluded.[2] Of the 57 SLE risk SNPs, 55 were available on the GSA array (table 1).

Of the 1,760 patients genotyped using the GSA, 576 did not pass the QC. We further excluded 44 individuals lacking clinical data or not meeting any of the ACR-82/ACR-97/SLICC-2012 classification criteria.[3-5] Moreover, 654 patients previously represented in the Immunochip dataset were removed to avoid duplication. Consequently, 486 patients from the GSA cohort remained for inclusion in the study, resulting in a combined total of 1,487 patients eligible for analysis.

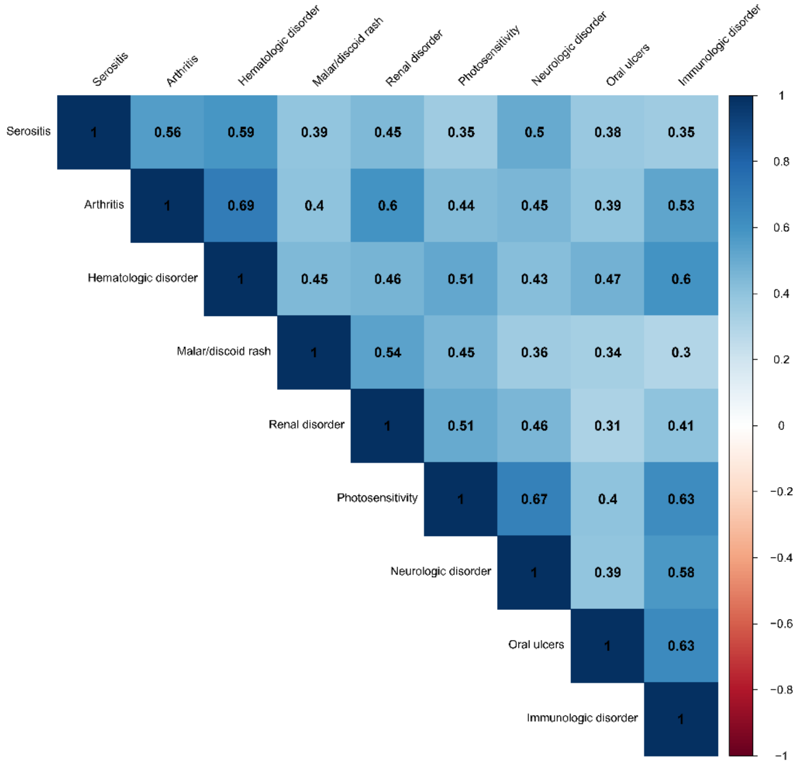


**Supplementary Figure 1: Correlation Matrix of ACR-82 Classification Criteria in SLE.** The matrix shows the analysis of SNP overlap between the different genetic risk scores (GRSs) used in the study.
